# Patterns of SARS-CoV-2 testing preferences in a national cohort in the United States

**DOI:** 10.1101/2020.12.22.20248747

**Authors:** Matthew L. Romo, Rebecca Zimba, Sarah Kulkarni, Amanda Berry, William You, Chloe Mirzayi, Drew Westmoreland, Angela M. Parcesepe, Levi Waldron, Madhura Rane, Shivani Kochhar, McKaylee Robertson, Andrew R. Maroko, Christian Grov, Denis Nash, for the CHASING COVID Cohort Study Team

## Abstract

In order to understand preferences about SARS-CoV-2 testing, we conducted a discrete choice experiment among 4793 participants in the Communities, Households, and SARS-CoV-2 Epidemiology (CHASING COVID) Cohort Study from July 30-September 8, 2020. We used latent class analysis to identify distinct patterns of preferences related to testing and conducted a simulation to predict testing uptake if additional testing scenarios were offered. Five distinct patterns of SARS-CoV-2 testing emerged. “Comprehensive testers” (18.9%) ranked specimen type as most important and favored less invasive specimen types, with saliva most preferred, and also ranked venue and result turnaround time as highly important, with preferences for home testing and fast result turnaround time. “Fast track testers” (26.0%) ranked result turnaround time as most important and favored immediate and same day turnaround time. “Dual testers” (18.5%) ranked test type as most important and preferred both antibody and viral tests. “Non-invasive dual testers” (33.0%) ranked specimen type and test type as similarly most important, preferring cheek swab specimen type and both antibody and viral tests. “Home testers” (3.6%) ranked venue as most important and favored home-based testing. By offering less invasive (saliva specimen type), dual testing (both viral and antibody tests), and at home testing scenarios in addition to standard testing scenarios, simulation models predicted that testing uptake would increase from 81.7% to 98.1%. We identified substantial differences in preferences for SARS-CoV-2 testing and found that offering additional testing options, which consider this heterogeneity, would likely increase testing uptake.

**SIGNIFICANCE:** During the COVID-19 pandemic, diagnostic testing has allowed for early detection of cases and implementation of measures to reduce community transmission of SARS-CoV-2 infection. Understanding individuals’ preferences about testing and the service models that deliver tests are relevant in efforts to increase and sustain uptake of SARS-CoV-2 testing, which, despite vaccine availability, will be required for the foreseeable future. We identified substantial differences in preferences for SARS-CoV-2 testing in a discrete choice experiment among a large national cohort of adults in the US. Offering additional testing options that account for or anticipate this heterogeneity in preferences (e.g., both viral and antibody tests, at home testing), would likely increase testing uptake.

**Classification:** Biological Sciences (major); Psychological and Cognitive Sciences (minor)

## INTRODUCTION

Diagnostic testing for SARS-CoV-2 infection has emerged as a critical tool in the public health armamentarium in the COVID-19 pandemic, as early detection allows for implementation of isolation and quarantine measures to reduce community transmission.^1^ The importance of testing has been well demonstrated globally, such as in South Korea, where a “test, trace, isolate” strategy has largely been credited for rapidly controlling transmission.^2^ Unfortunately, diagnostic testing in the US has been insufficient, which has led to both undetected cases transmitting disease in the community and an underestimation of the burden of COVID-19.^3^ Therefore, increasing testing uptake remains a major US public health priority in efforts to control community transmission as the pandemic continues to surge. Currently, the US Centers for Disease Control and Prevention recommends diagnostic testing for individuals with symptoms of COVID-19, individuals in close contact with a confirmed COVID-19 case, and other individuals at high risk of infection.^4^

Individuals’ preferences about testing, specifically about the test itself or the service model that delivers the test, are an important consideration in determining strategies to increase the uptake of SARS-CoV-2 testing. In other contexts, individual preferences about a health-related product or service have been shown to be predictive of health-related behaviors.^5^ Discrete choice experiments (DCEs), which are surveys that elicit stated preferences to identify trade-offs that a person makes with a good or service, have emerged as a tool to understand patient preferences and barriers to healthcare engagement,^6^ and are increasingly being used to inform patient centered healthcare.^7,8^ We previously conducted a DCE to understand SARS-CoV-2 testing preferences and found strong preferences for both viral and antibody testing, less invasive specimen collection, and rapid result turnaround time.^9^ However, as observed in DCEs on other topics, patient preferences are often heterogeneous and there may be distinct patterns of preferences within a sample.^10^

If indeed preferences are relevant to SARS-CoV-2 testing uptake and different patterns of preferences exist, these patterns may also be characterized by distinct demographic profiles. Previous work has documented demographic disparities in SARS-CoV-2 testing uptake. For example, among individuals receiving care at US Department of Veterans Affairs (VA) sites, SARS-CoV-2 testing rates within the VA system were lowest among whites, especially among those who were male and those who lived in rural settings.^11^ Patterns of preferences may also differ based on past experience with a product or service, as observed with frequency of past testing in preferences for HIV self-testing.^12^ Perceived concern or risk is another component involved in making decisions about health and since it is not uniformly distributed, it may also differ by patterns of preferences.^13^

We hypothesized that discernable patterns of SARS-CoV-2 testing preferences would emerge and that individuals in these patterns would have distinct demographic profiles, SARS-CoV-2 testing history, and perceived concern about infection.

## RESULTS

### Patterns of preferences: relative attribute importance and preferences for levels of attributes

Among the 4,793 participants who took the DCE, five distinct patterns of preferences were identified: *comprehensive testers* (18.9%), *fast track testers* (26.0%), *dual testers* (18.5%), *non-invasive dual testers* (33.0%), and *home testers* (3.6%). Each pattern had a distinct profile regarding the relative importance of attributes (Figure 1) and preferences for specific levels of attributes (Supplemental table 1):

**Figure 1.**
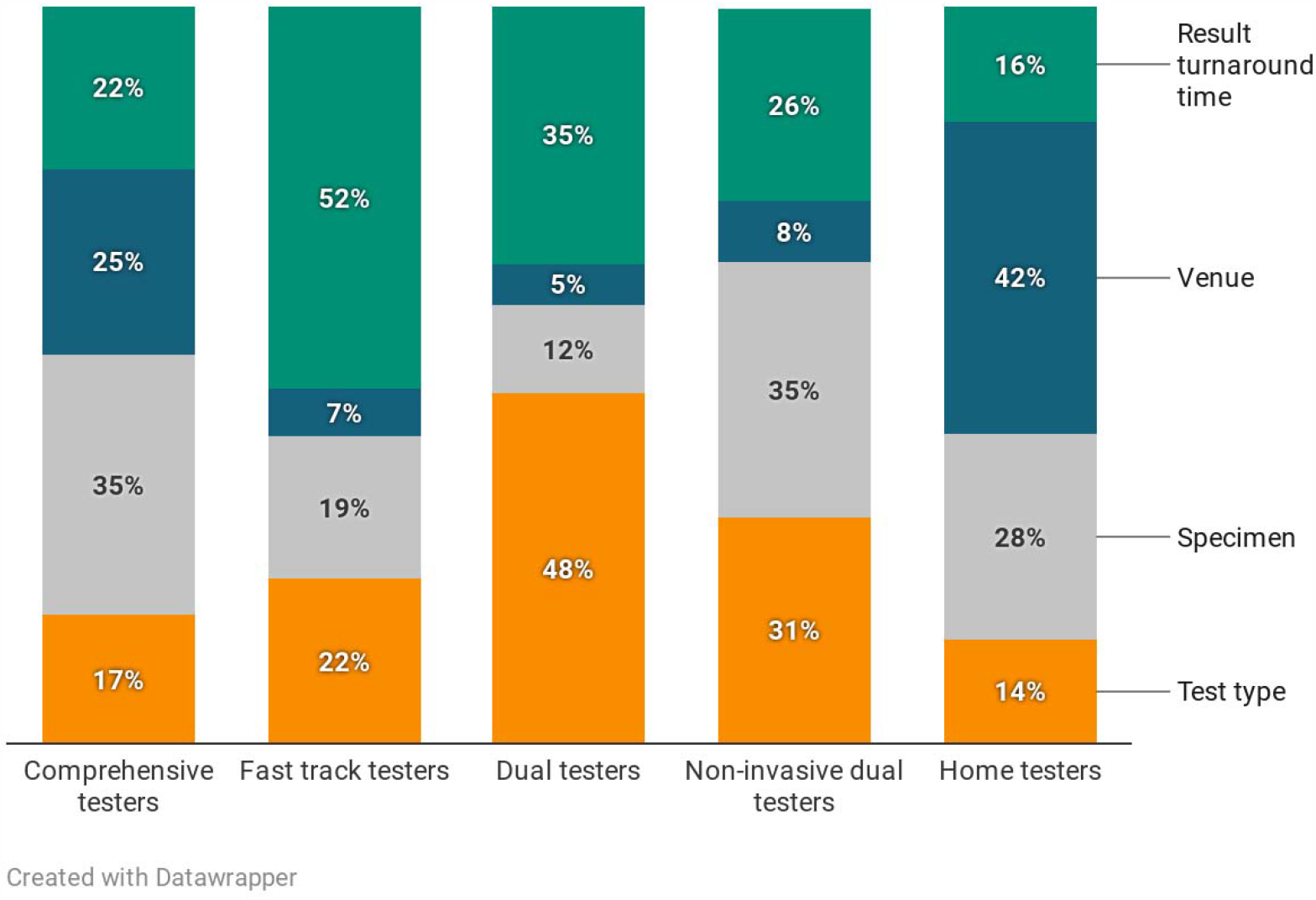
Mean relative attribute importance for SARS-CoV-2 testing by preference pattern.

**Figure 2.**
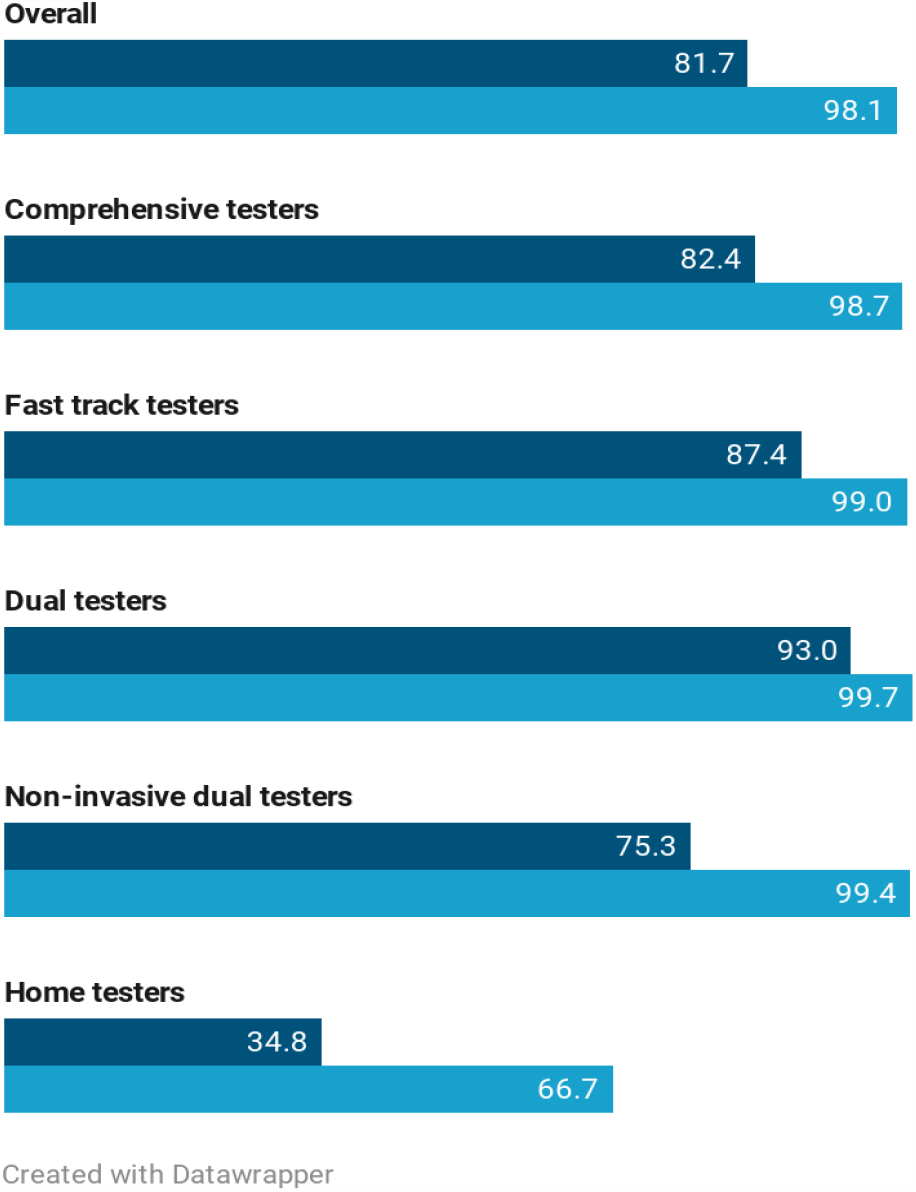
Predicted uptake of SARS-CoV-2 testing for two standard testing scenarios vs. the addition of less invasive dual testing, and at home scenarios, overall and by preference pattern.

*Comprehensive testers* ranked specimen type as most important (35%), followed by venue (25%), result turnaround time (23%), and test type (17%). Participants in this pattern least preferred nasopharyngeal swab and blood draw as specimen type (utilities -92.1 and - 41.5, respectively), and favored less invasive specimen types, with saliva most preferred (utility 48.8). Participants in this pattern preferred home sample collection, either returning the sample for testing by mail (utility 55.7) or to a collection site (utility 43.0), and least preferred testing at a doctor’s office/urgent care clinic (utility -41.9) or walk-in community testing site (utility -44.8). Participants in this pattern most preferred a fast turnaround time for results (immediate, utility: 41.7; same day, utility 30.2) and both antibody and viral tests (utility 39.3).

*Fast track testers* ranked result turnaround time as most important (52%), followed by test type (22%) and specimen type (19%); venue was least important (7%). Participants in this pattern most preferred immediate (utility 98.3) and same day (utility 63.7) result turnaround time and both antibody and viral tests (utility 53.0). Participants in this pattern preferred less invasive specimen types, with a cheek swab as most preferred (utility 25.4) and nasopharyngeal swab and blood draw as least preferred (utilities -51.4 and -20.7, respectively). Although venue was least important for this group, among the specific options, testing at a drive-thru community site was most preferred (utility 10.8).

*Dual testers* ranked test type as most important (48%), followed by result turnaround time (35%), with specimen type less important (12%) and venue least important (5%). Participants in this pattern most preferred both antibody and viral tests (utility 93.3) and preferred fast turnaround times for results (immediate, utility: 64.9; same day, utility 40.5). Less invasive specimen types were preferred, with check swab (utility 16.4) and saliva most preferred (utility 14.1) and nasopharyngeal swab least preferred (utility -30.4). Regarding venue, testing at drive-thru community sites was most preferred (utility 12.2).

*Non-invasive dual testers* ranked specimen type as most important (35%), closely followed by test type (31%) and then result turnaround time (26%), with venue least important (8%). Participants in this pattern most preferred specimen type was cheek swab (utility 37.9) as a specimen type and the least preferred nasopharyngeal swab (utility -100.9). Both antibody and viral tests were preferred (utility 73.1), as well as fast turnaround times for results (immediate, utility 54.6; same day, utility 25.0). Regarding venue, home collection with returning the sample to a collection site was most preferred (utility 17.1).

*Home testers* ranked venue as most important (42%), followed by specimen type (28%), and result turnaround time (16%) and test type (14%) were similarly less important. Participants in this pattern most preferred home sample collection either returning the sample for testing by mail (utility 93.3) or to a collection site (utility 60.3) and least preferred testing at a walk-in community site (utility -76.0) or a doctor’s office/urgent care (utility -44.6). Participants in this pattern in general preferred less invasive specimens and least preferred nasopharyngeal swab (utility -77.6) and blood draw (utility -21.0). Although these were least important attributes, participants with this pattern preferred both antibody and viral tests (utility 36.1) and fast turnaround times for results (immediate, utility: 32.4; same day, utility 21.2).

### Simulated preferences for standard testing, less invasive testing, dual testing, and at-home testing

In our simulation for standard testing, predicted testing uptake for the two scenarios overall was 81.7%, ranging from 34.8% for *home testers* to 93.0% for *dual testers* (Figure 1). By including less invasive testing, dual testing, and at home testing scenarios, predicted testing uptake increased by 16.4% to 98.1%. The addition of these three scenarios had the biggest impact on *home testers*, which increased from 34.8% to 66.7% uptake, and *non-invasive dual testers*, which increased from 75.3% to 99.4% uptake. In our simulation of all scenarios (Table 1, the standard testing scenarios generally had the lowest predicted uptake, with higher uptake for the drive-through option (overall 6.6%) compared with the walk-in community site option (overall 0.9%). Uptake for the standard testing scenarios was lowest for *non-invasive dual testers* (drive-through 1.3%, walk-in community site 0.1%). The dual testing scenario (of the three additional scenarios) had the highest predicted uptake overall (61.8%) and was highest for *fast track testers* (60.8%), *dual testers* (65.0%), and *non-invasive dual testers* (80.9%). The at-home testing scenario had the highest predicted uptake for *comprehensive testers* (38.0%) and *home testers* (38.1%); however, for *comprehensive testers*, there was also similar uptake for the dual testing scenario (35.4%), and for *home testers*, one-third (33.3%) did not have predicted uptake for any testing scenario.

**Table 1.**
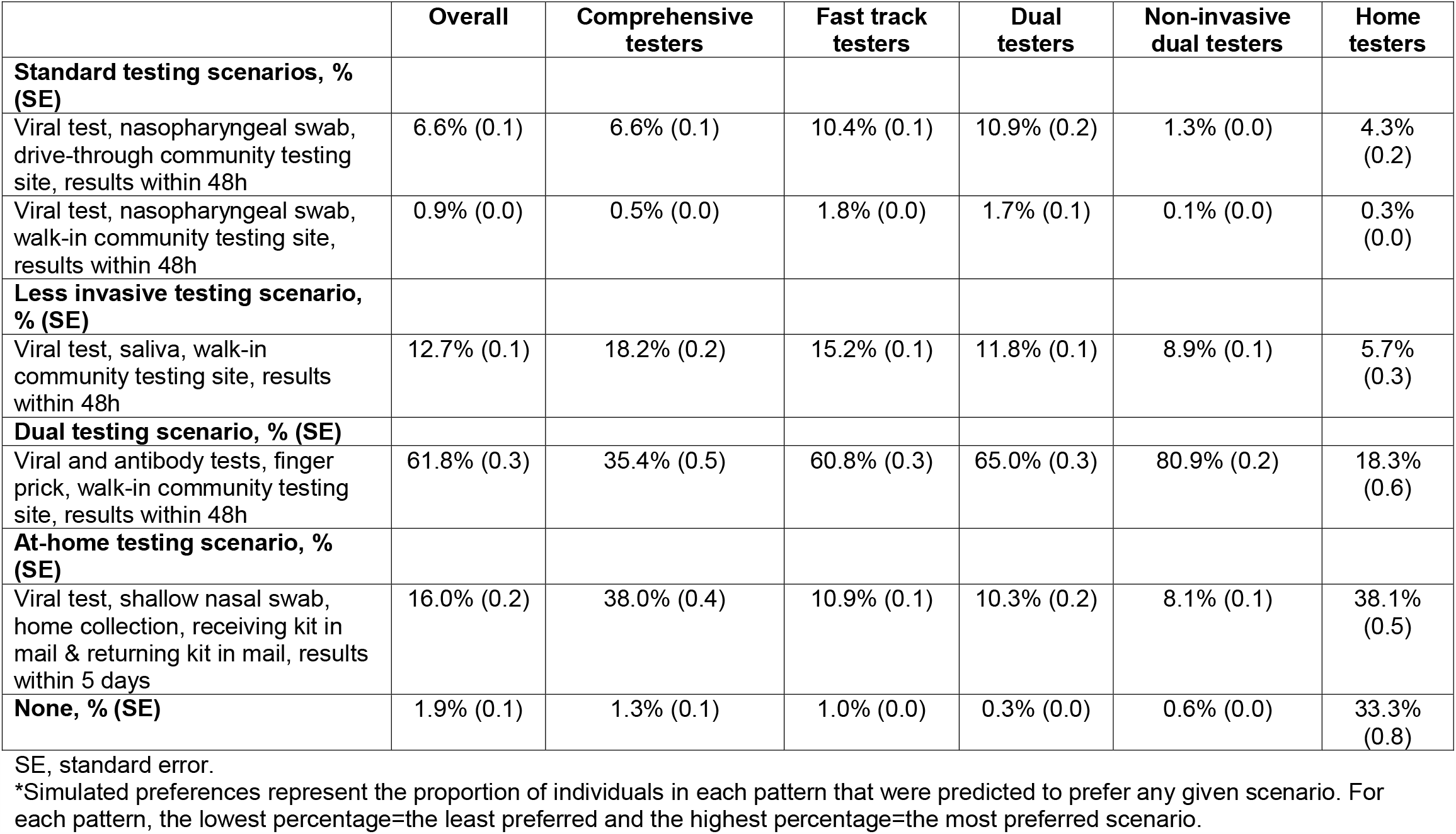
Simulated preferences* for standard testing, less invasive testing, dual testing, and at-home testing scenarios by preference pattern.

### Demographic characteristics of participants by preference pattern

There were statistically significant differences by preference pattern in age group, gender, race/ethnicity, education, and employment, but not region, urbanicity, or presence of any comorbidity (Table 2). *Home testers* were older and less often had representation in the youngest age group of 18-39 years (40.4%) compared with participants in other patterns (range 49.4%-56.0%). *Fast track testers* were less often female (47.3%), especially when compared with *home testers* (58.5%) and to a lesser extent when compared with participants in other patterns (range 50.3%-53.7%). *Dual testers* (70.4%) and *non-invasive dual testers* (67.1%) tended to be non-Hispanic white, especially compared with *home testers* (50.0%) and to a lesser extent with *comprehensive testers* (56.1%) and *fast track testers* (58.8%). *Dual testers* tended to be college graduates (70.9%), especially compared with *comprehensive testers* (55.2%) and to a lesser extent when compared with participants in other patterns (range 59.1%- 64.3%). Regarding employment, *home testers* were more often out of work (21.8%) compared with participants in other patterns (range 9.5%-13.9%).

**Table 2.** Sample characteristics, previous testing, COVID-19 diagnosis, and concern about infection stratified by preference pattern for SARS-CoV-2 testing.

|  | **Comprehensive testers** | **Fast track testers** | **Dual testers** | **Non-invasive dual testers** | **Home testers** | **p-value** |
| --- | --- | --- | --- | --- | --- | --- |
|  | *N* = 907 | *N* = 1245 | *N* = 889 | *N* = 1581 | *N* = 171 |  |
| **Age group, *n* (%)** |  |  |  |  |  | .002 |
| 18-39 years | 508 (56.0%) | 667 (53.6%) | 464 (52.2%) | 781 (49.4%) | 69 (40.4%) |  |
| 40-59 years | 269 (29.7%) | 382 (30.7%) | 280 (31.5%) | 503 (31.8%) | 68 (39.8%) |  |
| ≥60 years | 130 (14.3%) | 196 (15.7%) | 145 (16.3%) | 297 (18.8%) | 34 (19.9%) |  |
| **Gender, *n* (%)** |  |  |  |  |  | .011 |
| Female | 456 (50.3%) | 589 (47.3%) | 474 (53.3%) | 849 (53.7%) | 100 (58.5%) |  |
| Male | 428 (47.2%) | 623 (50.0%) | 385 (43.3%) | 691 (43.7%) | 66 (38.6%) |  |
| Transgender, non-binary, other | 23 (2.5%) | 33 (2.7%) | 30 (3.4%) | 41 (2.6%) | 5 (2.9%) |  |
| **Race/ethnicity, *n* (%)** |  |  |  |  |  | <.001 |
| Hispanic | 177 (19.6%) | 241 (19.4%) | 117 (13.2%) | 218 (13.8%) | 35 (20.6%) |  |
| Non-Hispanic Black | 114 (12.6%) | 128 (10.3%) | 68 (7.7%) | 136 (8.6%) | 25 (14.7%) |  |
| Asian/Pacific Islander | 66 (7.3%) | 96 (7.7%) | 63 (7.1%) | 118 (7.5%) | 13 (7.7%) |  |
| Non-Hispanic white | 508 (56.1%) | 732 (58.8%) | 625 (70.4%) | 1059 (67.1%) | 85 (50.0%) |  |
| Other | 40 (4.4%) | 47 (3.8%) | 15 (1.7%) | 47 (3.0%) | 12 (7.1%) |  |
| **Education, *n* (%)** |  |  |  |  |  | <.001 |
| Less than high school | 15 (1.7%) | 17 (1.4%) | 10 (1.1%) | 25 (1.6%) | 2 (1.2%) |  |
| High school graduate | 120 (13.3%) | 107 (8.6%) | 64 (7.2%) | 133 (8.4%) | 24 (14.0%) |  |
| Some college or technical school | 271 (29.9%) | 337 (27.1%) | 184 (20.8%) | 405 (25.7%) | 44 (24.7%) |  |
| College graduate | 500 (55.2%) | 783 (62.9%) | 628 (70.9%) | 1015 (64.3%) | 101 (59.1%) |  |
| **Employment, *n* (%)** |  |  |  |  |  | <.001 |
| Employed | 589 (65.0%) | 812 (65.3%) | 579 (65.1%) | 1008 (63.8%) | 95 (55.9%) |  |
| Out of work | 118 (13.0%) | 173 (13.9%) | 84 (9.5%) | 194 (12.3%) | 37 (21.8%) |  |
| Not in the workforce | 199 (22.0%) | 259 (20.8%) | 226 (25.4%) | 379 (24.0%) | 38 (22.4%) |  |
| **Region, *n* (%)** |  |  |  |  |  | .122 |
| Northeast | 200 (22.3%) | 312 (25.2%) | 243 (27.6%) | 388 (24.8%) | 46 (27.2%) |  |
| Midwest | 141 (15.7%) | 229 (18.5%) | 160 (18.1%) | 298 (19.0%) | 22 (13.0%) |  |
| South | 317 (35.3%) | 409 (33.0%) | 282 (32.0%) | 489 (31.3%) | 63 (37.3%) |  |
| West | 237 (26.4%) | 287 (23.2%) | 196 (22.2%) | 388 (24.8%) | 38 (22.5%) |  |
| US Dependent | 3 (0.3%) | 1 (0.1%) | 1 (0.1%) | 2 (0.1%) | 0 (0.0%) |  |
| **Urban, *n* (%)** |  |  |  |  |  | .308 |
|  | 371 (40.9%) | 560 (45.0%) | 398 (44.8%) | 673 (42.6%) | 72 (42.1%) |  |
| **Any comorbidity, *n* (%)** |  |  |  |  |  | .553 |
|  | 349 (38.5%) | 495 (39.8%) | 343 (38.6%) | 631 (39.9%) | 77 (45.0%) |  |
| **Previously tested for SARS-CoV-2, *n* (%)** |  |  |  |  |  | <.001 |
|  | 236 (26.0%) | 428 (34.5%) | 298 (33.5%) | 378 (23.9%) | 40 (23.4%) |  |
| **Previously received laboratory-confirmed diagnosis of COVID-19, *n* (%)** |  |  |  |  |  | <.001 |
|  | 56 (6.2%) | 94 (7.6%) | 35 (3.9%) | 51 (3.2%) | 9 (5.3%) |  |
| **Concern about getting infected with SARS-CoV-2 (among those without COVID-19 diagnosis), *n* (%)** |  |  |  |  |  | <.001 |
| Not at all worried/not too worried | 229 (26.9%) | 277 (24.1%) | 180 (21.1%) | 455 (29.7%) | 52 (32.1%) |  |
| Somewhat worried | 403 (47.4%) | 576 (50.0%) | 472 (55.3%) | 760 (49.7%) | 68 (42.0%) |  |
| Very worried | 219 (25.7%) | 298 (25.9%) | 202 (23.7%) | 315 (20.6%) | 42 (25.9%) |  |
| **Concern about loved ones getting infected with SARS-CoV-2, *n* (%)** |  |  |  |  |  | <.001 |
| Not at all worried/not too worried | 160 (17.7%) | 161 (12.9%) | 82 (9.2%) | 256 (16.2%) | 38 (22.4%) |  |
| Somewhat worried | 331 (36.5%) | 529 (42.5%) | 387 (43.5%) | 658 (41.6%) | 65 (38.2%) |  |
| Very worried | 415 (45.8%) | 555 (44.6%) | 420 (47.2%) | 667 (42.2%) | 67 (39.4%) |  |

### Previous SARS-CoV-2 testing, COVID-19 diagnosis, and infection concern by preference pattern

There were statistically significant differences by preference pattern for previous SARS-CoV-2 testing, reported COVID-19 diagnosis, concern about getting infected, and concern about loved ones getting infected (Table 2). *Fast track testers* (34.5%) and *dual testers* (33.5%) more often had previously tested compared with participants in other patterns (range 23.4%-26.0%). Among all participants, *dual testers* (3.9%) and *non-invasive dual testers* (3.2%) less often had a previous laboratory-confirmed diagnosis of COVID-19 than participants in other patterns (range 5.3%-7.6%). *Dual testers* were less often (21.1%) not at all worried/not too worried about getting infected with SARS-CoV-2, especially when compared with *home testers* (32.1%) and to a lesser extent, participants in other patterns (range 24.1%-29.7%). A similar pattern was observed for concern about loved ones getting infected with SARS-CoV-2, with *dual testers* less often not at all worried/not too worried about loved ones getting infected (9.2%) compared with *home testers* (22.4%) and to a lesser extent compared with participants in other patterns (range 12.9%-17.7%).

## DISCUSSION

We identified substantial differences in preferences for SARS-CoV-2 testing, as shown by the differences in the relative importance of test type, specimen, venue, and result turnaround time. For example, test type was very important for participants in some patterns, specifically *dual testers* and *non-invasive dual testers*, whereas this was the least important attribute for *comprehensive testers* and *home testers*. Regarding attribute levels, we observed some commonalities across all patterns—for example, participants preferred getting both antibody and viral tests with a less-invasive specimen and fast turnaround time for results.

However, there were substantial differences in preferences for venue of testing. In addition to venue being highly important for *comprehensive testers* and *home testers*, home testing was strongly preferred among those participants. Venue was least important for *fast track testers, dual testers*, and *non-invasive dual testers* who had varying preferences for venue types. Among *fast track testers* and *dual testers*, community-based drive-through testing was favored. Our simulation showed that offering additional options would increase testing uptake, with dual viral and antibody testing expected to have the biggest effect on testing uptake. An at-home testing option would also be expected to increase uptake, especially for participants in *comprehensive testers* and *home testers*, which consisted of about one-fifth of the sample.

We also observed differences in sample characteristics by preference pattern, including demographic characteristics, previous testing and COVID-19 diagnosis, and concern about infection. For example, *dual testers*, who placed most importance on having both viral and antibody testing, were disproportionately white and college graduates and were most concerned about infection for themselves and loved ones compared with participants in other patterns. Although they were more likely than participants in other patterns to have previously tested, they were less likely to have a previous COVID-19 diagnosis. In contrast, *comprehensive testers* and *home testers*, who placed most importance on specimen and venue, respectively, were more often Black and Hispanic and non-college graduates and were less concerned about infection for themselves and loved ones. These differences in preference patterns by race may provide insight on how to increase testing among Black and Latino individuals, who experience a disproportionate burden of cases, hospitalization, and deaths due to COVID-19.^14^ We did not find a significant difference in pattern of preferences by urbanicity, which was unexpected because individuals in non-urban, rural areas have longer travel times to testing sites^15^ and we would anticipate they would be more likely to have preferences for home testing. Relatedly, we did not observe a significant difference in preference pattern by aggregate geographic region, which also was unexpected because we would anticipate that differences in local outbreak patterns and local health departments responses may impact preferences about testing.

Our results should be interpreted in the context of their limitations. First, an important limitation of our analysis is related to latent class analysis in general, as best practices for using it to study heterogeneity in preferences in health-related research are still evolving.^10^ For example, regarding number of classes, for which there is no consensus, we selected 5, after comparing sample size, fit statistics, and overall interpretability of 2-10 classes. It is therefore possible that there were some patterns of preferences that went unidentified. Second, participants’ preferences about SARS-CoV-2 testing may change over time. Research on other topics has demonstrated that choices stated in a DCE are generally consistent, with good test-retest reliability;^16,17^ however, knowledge about SARS-CoV-2 and COVID-19 is rapidly evolving and is widely disseminated in mainstream media,^18^ which could plausibly impact preferences. For example, reports of re-infection and the potential waning of antibodies, such as the first such report in the US in October 2020,^19^ approximately one month after the completion of our DCE, might influence preferences about antibody testing, as might the availability of highly efficacious vaccines in December 2020.^20^ Third, stated preferences regarding SARS-CoV-2 testing in our DCE may not necessarily align with actual behavior (i.e., revealed preferences); however, a systematic review and meta-analysis found that, in general, stated preferences in DCEs aligned with revealed preferences.^5^ Reasons for lack of concordance between stated and revealed preferences include omitting attributes in DCEs that may present in real-life decisions, such as aspects of accessibility including cost, transportation time, availability of testing, and wait time. Therefore, future work on external validation of stated preference research relevant to COVID-19 is warranted. Fourth, although our sample was large and geographically diverse, it was not a representative sample, so it may be that there are additional patterns of preferences that exist beyond our study. Furthermore, most participants (78.3%) in the DCE had already completed at-home self-collection of a dried blood spot specimen as part of our larger cohort study, which may have influenced preferences regarding the venue of testing. Finally, it will be important to examine preferences as new testing modalities become available, such as a new fully at-home test that provides rapid results,^21^ and as vaccine uptake increases.

Our study may inform ways to better design and deliver SARS-CoV-2 testing services in line with pandemic response goals. The heterogeneity in preferences observed by pattern highlights that having more options available (and educating the public about their availability) is one way to increase testing uptake. For example, coupling antibody and viral testing (e.g., offering antibody testing to individuals following negative PCR results) is a way that health departments could potentially increase uptake. Home-based testing could also increase uptake, especially for individuals who place importance on venue of testing, so it should also be considered in testing programs. Importantly, our analysis highlights that preferences for SARS-CoV-2 testing differ by population characteristics, including demographics, which must also be considered in the context of existing health disparities in the US. Even as highly effective vaccines for SARS-CoV-2 become available, we anticipate that testing will remain a critical tool in the pandemic response until vaccine coverage and herd immunity are sufficiently high enough to reduce transmission; thus, understanding preferences for SARS-CoV-2 testing will remain an important endeavor.

## MATERIALS AND METHODS

The survey design of our DCE has been previously described,^9^ but here we provide a summary. Participants in the Communities, Households, and SARS-CoV-2 Epidemiology (CHASING COVID) Cohort Study^22^ who completed a follow-up assessment (*N* = 5,098) were invited to complete the DCE. All study procedures were approved by the CUNY Graduate School of Public Health and Health Policy Institutional Review Board. The DCE was designed and implemented using Lighthouse Studio 9.8.1 (Sawtooth Software, Provo, UT) and each participant was presented with 5 choice tasks where they were asked to indicate which of two different testing scenarios was preferable or if neither was acceptable. Specific testing attributes (and levels) examined were type of test (antibody, viral, or both), specimen type (finger prick, blood draw, check swab, saliva, shallow nasal swab, nasopharyngeal swab, and urine), testing venue (home collection, receiving kit in mail & returning kit in mail; home collection, receiving kit in mail & returning to a collection site; doctor’s office or urgent care clinic; walk-in community testing site; drive-thru community testing site; pharmacy), and result turnaround time (immediate, same day, 48 hours, 5 days, >5 days). Of the 5,098 participants in the cohort invited, 4,793 participants (94.0%) completed the DCE from July 30 through September 8, 2020. The median age was 39 years, 51.5% were female, 62.8% were non-Hispanic white, 16.5% were Hispanic, 9.8% were non-Hispanic Black, and 7.1% were Asian.^9^ At enrollment, 29.3% of participants resided in the Northeast, 28.6% in the South, 24.1% in the West, and 17.9% in the Midwest.

Other measures of interest were collected from participants in the CHASING COVID Cohort Study^22^ during the baseline survey (age, gender, race/ethnicity, education, region, urbanicity, comorbidities) and a combination of baseline and follow-up surveys (employment, concern about infection, previous SARS-CoV-2 testing). For this analysis, age was categorized into 3 groups (18-39 years, 40-59 years, and ≥60 years); gender was recategorized into 3 groups (male, female, transgender/non-binary/other); race/ethnicity was ascertained by two questions related to Hispanic heritage and race, and was categorized into 5 groups (Hispanic, non-Hispanic Black, Asian/Pacific Islander, non-Hispanic white, and other); education was ascertained by asking about highest grade or year of school completed (less than high school diploma, grade 12 or GED [high school graduate], college 1-3 years [some college or technical school], college 4 years or more [college graduate]); region was ascertained based on US state or territory of residence and categorized into 4 census regions (Northeast, Midwest, South, West) and Puerto Rico; urbanicity was categorized based on city locale assignments from the National Center for Education Statistics using ZIP Code Tabulation Areas with urban defined as living in an urbanized area and inside a principal city;^23^ and having any comorbidities (yes/no) was based on a question about ever being told by a healthcare professional that the participant had any chronic conditions in a list (i.e., coronary artery disease, diabetes, hypertension, cancer, asthma, chronic obstructive pulmonary disease, kidney disease, HIV/AIDS, immunosuppression, and depression). Employment was ascertained in each survey by asking about the participant’s current situation and was recategorized as employed (employed for wages, self-employed), out of work, or not in the workforce (homemaker, student, retired). Participants were asked in baseline and follow-up surveys about past SARS-CoV-2 testing and frequency of testing, which we recategorized as never (inclusive of don’t know/not sure) or yes (one or more times). Participants were also asked in the baseline and follow-up surveys if they had previously received a laboratory-confirmed diagnosis of COVID-19. For concern about infection, participants were asked separate questions about the intensity of their concern about themselves or loved ones getting sick from SARS-CoV-2 with responses recategorized as not at all worried/not too worried, somewhat worried, and very worried. The question regarding concern about themselves getting infected was restricted to participants who reported not having a previous laboratory-confirmed diagnosis of COVID-19.

We used latent class analysis to determine mean relative attribute importance and zero-centered part-worth utilities for each attribute level within each pattern. We ran the latent class analysis with 2-10 classes; however, to maximize both sample size within groups and interpretability and after examination of fit statistics (e.g., Bayesian information criterion), we present data for 5 classes that we interpret as distinct patterns of preferences. Latent class estimation first selected random estimates of each patten’s utility values, then used these values to fit each participant’s data and estimate the relative probability of each respondent belonging to each pattern.^24^ Next, using probabilities as weights, logit weights were re-estimated for each pattern and log-likelihoods were accumulated across all patterns. This process was repeated until reaching the convergence limit.

We then simulated preferences within each of the 5 preference patterns for 5 testing approaches:

1 and 2) “Standard testing” was based on major health departments’ current testing programs with viral test (nasopharyngeal swab) and result turnaround time of 48 hours, and consisted of two unique scenarios because of venue:^25,26^ drive-through testing site and walk-in community testing site. Result turnaround time was based on reports from major health departments (e.g., >85% of results within 2 days in California).^25^

3) “Less invasive testing” was based on some jurisdictions offering less invasive specimen collection, such as saliva:^25^ viral test, saliva specimen, walk-in community testing site, and result turnaround time of 48 hours.

4) “Dual testing” consisted of both viral and antibody testing and would necessitate a finger prick:^27^ viral and antibody test, finger prick specimen, walk-in community testing site, and result turnaround time of 48 hours.

5) “At-home testing” was based on a commercially available at-home testing kit:^28^ viral test, shallow nasal swab, home collection, receiving kit in mail & returning kit in mail, and result turnaround time of within 5 days, as additional time would be required for mailing the specimen.

We conducted two simulations: 1) the two standard testing scenarios with a “no test” option to capture the proportion of participants in each pattern who would opt out of testing altogether, given the choices; 2) the two standard test scenarios, less invasive, dual testing, and at home testing scenarios, and a “no test” option. Preferences for the 3 total options for the first simulation and 6 total options for the second simulation were generated using randomized first choice models whereby utilities were summed across all levels for each scenario and then exponentiated and rescaled to sum to 100.^29^

Finally, we computed descriptive statistics (frequencies and proportions) for demographic characteristics, previous SARS-CoV-2 testing, previous COVID-19 diagnosis, and concern about infection stratified by the 5 preference patterns and compared distributions of these variables using Pearson’s Chi-Square tests. An alpha level of 0.05 was the criterion for statistical significance. The latent class analysis was done using Lighthouse Studio 9.8.1 (Sawtooth Software, Provo, UT) and the descriptive statistics and bivariate analysis were done using SAS 9.4 (Cary, NC).

## Data Availability

De-identified data are available upon request from the corresponding author.

## DATA AVAILABILITY

De-identified data are available upon request from the corresponding author.

## ACKNOWLEGEMENTS

We would like to acknowledge the CHASING COVID Cohort Study participants for their contributions to this research.

---

**Supplemental table 1.** Zero-centered part-worth utilities* for SARS-CoV-2 testing attribute levels by preference pattern.

|  | **Comprehensive testers** | **Fast track testers** | **Dual testers** | **Non-invasive dual testers** | **Home testers** |
| --- | --- | --- | --- | --- | --- |
| **N, %** | 907 (18.9%) | 1245 (26.0%) | 889 (18.5%) | 1581 (33.0%) | 171 (3.6%) |
| **Test type, mean utility (SE)** |  |  |  |  |  |
| Antibody | -28.2 (0.5) | -34.4 (0.5) | -97.1 (0.5) | -26.4 (0.3) | -20.0 (0.3) |
| Viral | -11.1 (0.4) | -18.6 (0.3) | 3.7 (0.3) | -46.7 (0.2) | -16.1 (0.3) |
| Both tests | 39.3 (0.6) | 53.0 (0.3) | 93.3 (0.2) | 73.1 (0.2) | 36.1 (0.4) |
| **Specimen type, mean utility (SE)** |  |  |  |  |  |
| Finger prick | 23.5 (0.1) | 19.7 (0.1) | 2.0 (0.2) | 25.7 (0.1) | 24.5 (0.1) |
| Blood draw | -41.5 (0.4) | -20.7 (0.2) | -18.7 (0.1) | -1.7 (0.2) | -21.0 (0.3) |
| Cheek swab | 34.0 (0.1) | 25.4 (0.1) | 16.4 (0.2) | 37.9 (0.1) | 25.6 (0.1) |
| Saliva | 48.8 (0.4) | 13.4 (0.2) | 14.1 (0.2) | 28.1 (0.1) | 18.7 (0.3) |
| Shallow nasal swab | 9.4 (0.2) | 7.4 (0.1) | 6.7 (0.1) | -10.5 (0.1) | -3.5 (0.2) |
| Nasopharyngeal swab | -92.1 (0.5) | -51.4 (0.4) | -30.4 (0.5) | -100.9 (0.3) | -77.6 (0.4) |
| Urine | 18.0 (0.2) | 6.1 (0.1) | 9.9 (0.1) | 21.4 (0.1) | 33.5 (0.3) |
| **Venue, mean utility (SE)** |  |  |  |  |  |
| Home collection, receiving kit in mail & returning kit in mail | 55.7 (0.7) | 5.5 (0.2) | -4.1 (0.1) | 8.9 (0.2) | 93.3 (1.0) |
| Home collection, receiving kit in the mail & returning to a collection site | 43.0 (0.5) | -8.6 (0.2) | -3.7 (0.2) | 17.1 (0.2) | 60.3 (0.7) |
| Doctor's office or urgent care clinic | -41.9 (0.4) | -5.5 (0.1) | -4.0 (0.1) | -13.3 (0.1) | -44.6 (0.4) |
| Walk-in community testing site | -44.8 (0.4) | -6.7 (0.2) | -3.7 (0.2) | -16.2 (0.1) | -76.0 (0.8) |
| Drive through community testing site | 2.7 (0.1) | 10.8 (0.1) | 12.2 (0.1) | -3.6 (0.1) | -6.9 (0.2) |
| Pharmacy | -14.7 (0.3) | 4.5 (0.1) | 4.9 (0.1) | 7.2 (0.1) | -26.1 (0.4) |
| **Result turnaround time, mean utility (SE)** |  |  |  |  |  |
| Immediate | 41.7 (0.5) | 98.3 (0.3) | 64.9 (0.2) | 54.6 (0.2) | 32.4 (0.7) |
| Same day | 30.2 (0.3) | 63.7 (0.3) | 40.5 (0.2) | 25.0 (0.2) | 21.2 (0.5) |
| 48 hours | 1.3 (0.1) | 13.6 (0.1) | 8.4 (0.0) | 9.5 (0.0) | -5.2 (0.2) |
| 5 days | -25.9 (0.3) | -66.8 (0.2) | -39.0 (0.2) | -39.1 (0.1) | -18.5 (0.5) |
| Greater than 5 days | -47.3 (0.5) | -108.8 (0.4) | -74.8 (0.3) | -50.0 (0.3) | -30.0 (0.8) |
| **None, mean utility (SE)** | -299.7 (1.2) | -238.3 (0.5) | -217.3 (0.5) | -226.2 (0.4) | 32.4 (4.4) |

SE, standard error.
\*Direction of the utility values implies the preference for the attribute level (negative=less preferred; positive=more preferred) and the magnitude of the utility implies the strength of preference.

